# Estimating future vaccine manufacturing capacity requirements using discrete event simulation: a case study on Africa’s 2040 vision

**DOI:** 10.64898/2026.07.05.26357327

**Authors:** Junior Ocira, Donovan Guttieres, Robin Kelchtermans, Eki Ramnaps Eghosasere, Carla Van Riet, Lise Boey, Kristopher Howard, Mauro Bernuzzi, Steven D. Dong, Jens Demand, Simone Blayer, Nico Vandaele

## Abstract

The Africa CDC’s New Public Health Order aims to improve health, regional self-reliance, and health security by boosting local vaccine manufacturing capacity from <1% in 2022 to 60% by 2040. Although multiple studies have examined vaccine manufacturing capacity in Africa, none quantified future vaccine manufacturing capacity requirements and complexities in capacity planning. We fill this gap by integrating a deterministic approach and discrete event simulation (DES) to estimate vaccine manufacturing capacity requirements and assess the impact of process variability and uncertainty on manufacturing capacity needs. A total of 51 experts were interviewed to design vaccine-specific supply chain networks, collect, and validate data on process-specific parameters. Our findings reveal that deterministic approach provides optimistic capacity estimates but realistic estimates of capacity requirements is possible using DES as it captures variability and uncertainty inherent in vaccine manufacturing. Further evidence shows that reduction in batch yield at any manufacturing stage significantly reduces throughput, while the impact of high batch deviations is vaccine specific and dependent on the manufacturing stage. For multi-antigen vaccines, capacity estimation is complicated by the amplification of process variability, and synchronization of batch arrival and volume. Finally, we estimated that 11 vaccine manufacturing facilities staffed by approximately 8,599 fulltime employees, of whom 43% are technical personnel, would be required to achieve Africa’s 2040 ambition. Overall, adopting a system-level perspective, maintaining a high, reliable, and sustainable manufacturing process yield and quality, are important when establishing new vaccine manufacturing facilities.

## Introduction

The Africa CDC (AfCDC) estimates that vaccine demand in Africa will double between 2022 and 2040, reaching approximately 2.7 billion doses. As of 2022, limited vaccine manufacturing capacity existed in Africa, with less than 1% of demand being produced locally ^1^. Hence, AfCDC has set a bold vision to manufacture 60% of African vaccine demand locally by 2040 ^1^. To reach this 2040 ambition, the AfCDC prioritised several vaccines targeting 22 diseases for fill-and-finish (F&F) production, while high-volume vaccines and those targeting African-endemic diseases were prioritised for both integrated drug substance (DS) and F&F production locally. These prioritized vaccines are: Pentavalent, Measles and Rubella (MR), Malaria, Human Immunodeficiency Virus (HIV), Pneumococcal Conjugate Vaccine (PCV), Bacillus Calmette-Guérin (BCG), Yellow fever, COVID-19, Outbreak (Ebola, Influenza, Chikungunya, Rift Valley fever, Lassa fever, Disease X) ^1^.

In terms of locally integrated DS and F&F production capacity, only two manufacturers have the manufacturing capacity in Africa as of 2022: the Institut Pasteur de Dakar has capacity to produce 5 to 8 million doses of Yellow fever vaccine annually ^2^, and the Institut Pasteur de Tunis produced about 18,680 doses of BCG vaccine in 2022 ^3^. To determine the vaccine manufacturing capacity that still needs to be installed in Africa, the AfCDC estimated that 15 new vaccine manufacturing facilities with about 6,000 new fulltime employees (FTEs) are required to produce around 1.6 billion doses annually by 2040^1^. These estimates were based on consultations with industry experts on the compatibility of different vaccine manufacturing technologies, the minimum economically viable and average size of new vaccine manufacturing facilities, and data on the number of equipment units, personnel shifts, and full-time equivalents (FTEs) per shift for DS and F&F manufacturing across different vaccine technologies ^1^.

Estimating vaccine manufacturing capacity is challenging because of the complexities inherent in the manufacturing processes. These complexities include yield uncertainties, long and variable lead times, complex quality assurance and quality control (QA/QC) procedures, inevitable equipment breakdowns, and extended maintenance periods ^4,5^. Despite these complexities and uncertainties, vaccine manufacturers must make early investment decisions regarding how much manufacturing capacity to install ^6^. Uncertain production batch yield, for example, has been shown to result in lower-than-expected production outputs ^7^, and impact the availability of seasonal influenza vaccines ^8^, malaria vaccines ^9^, and COVID-19 vaccines ^10^. In addition, factors like batch processing times, human resource planning, equipment setup and cleaning times, and QA/QC testing impacts overall capacity because they influence how many equipment units are required to meet demand within a given timeframe ^11,12^. These uncertainties can complicate capacity planning by having either (i) excess capacity leading to excess inventory or low utilisation eroding profitability, or (ii) under capacity leading to stock outs and/or long lead times, thus delaying access ^13^.

Simulation models have been used to assess how uncertainties influences vaccine manufacturing performance. For example, techno-economic approach has been used to estimate resources and capacity requirements for producing COVID-19 vaccines using different vaccine manufacturing technologies ^10,14^. Although this approach provide detailed capacity requirements including costs, they require granular input data not readily available in the publicly domain, especially for non-pandemic vaccines. Another simulation approach is discrete event simulation (DES) extensively used to design, analyse and evaluate complex manufacturing production systems, operational plans, and scheduling^15^. DES also enables uncertainties in manufacturing demand, supply, and equipment availability to be taken in to account ^16^. One study developed a generic DES model to explore how disruptions across the vaccine supply chain influence annual throughput, lead times, and resilience at an aggregate level ^17^.

Existing studies on vaccine manufacturing capacity in Africa focus on the following topics: feasibility of vaccine manufacturing business models ^18^, challenges faced in vaccine manufacturing scaleup ^19^, ecosystems approach towards establishing manufacturing capacity in Africa ^20^, and leveraging innovations, technologies, partnerships, and research and development for sustainable vaccine manufacturing ^21^. While these studies provide important strategic policy directions, they do not provide a structured flexible approach to estimating vaccine manufacturing capacity requirement to meet a target demand. In addition, there is limited evidence on how vaccine manufacturing process variabilities and uncertainties impacts capacity requirements for different vaccine manufacturing networks. This study addresses these gaps by combining deterministic capacity estimation approach with DES. The deterministic dimension provides a rapid approximation of the manufacturing capacity required given a target demand and most likely values of manufacturing process parameters ^22^. However, since it uses fixed parameter values, DES provides an alternative to incorporate manufacturing complexities, variabilities, and uncertainties, in addition to assessing their impact on capacity requirements and overall performance of manufacturing systems ^23^.

In this paper, we apply the DES model developed by Kelchtermans et al. ^17^ to determine the vaccine manufacturing capacity needed to fulfil demand for vaccines prioritised for integrated DS and F&F manufacturing in Africa by 2040. Through scenario analysis, we also show how changes in batch yield, batch size, processing times, equipment availabilities, quality deviations, and transportation delay impacts the estimated capacity.

## Methodology

Manufacturing capacity is multifaceted and consists of different aspects including available space, equipment, employees, input materials, or combination of these ^24^. In this research, we define manufacturing capacity as the total equipment units, annual throughput, total manufacturing facilities, and total full-time employees (FTEs) required to meet a target demand. An equipment unit used to process vaccine batches in DS can be bioreactor, IVT reactor, fixed bed reactor, conjugation reactor, fermenter, cell factories, roller bottles, scale X, shake flasks, or eggs. For F&F, the equipment units can be formulation vessel, filling line, lyophilizers, visual inspection line, or packaging line. Our framework (see Figure 1) starts by estimating the theoretical number of equipment units required for each manufacturing process given the vaccine manufacturing process parameters, vaccine manufacturing supply chain (VMSC) network, and annual demand. These are then used as inputs in the DES model to simulate a typical vaccine manufacturing process ^17^ shown in Figure 2. We then calculated key performance indicators (KPIs) and use it to evaluate whether the simulation average throughput meets the target vaccine demand. If the simulation average throughput is lower than demand, the bottleneck process is identified as the process with the highest average annual equipment utilisation, and the total of equipment units for that process is increased by one unit ^25^. We subsequently reran the DES model and the KPIs are recalculated and evaluated until the simulated average throughput exceed the target demand. The resulting total equipment units are integrated with operational staffing and facility sizing information, to calculate the total FTEs required. Detailed information follows in the next subsections.

**Figure 1:**
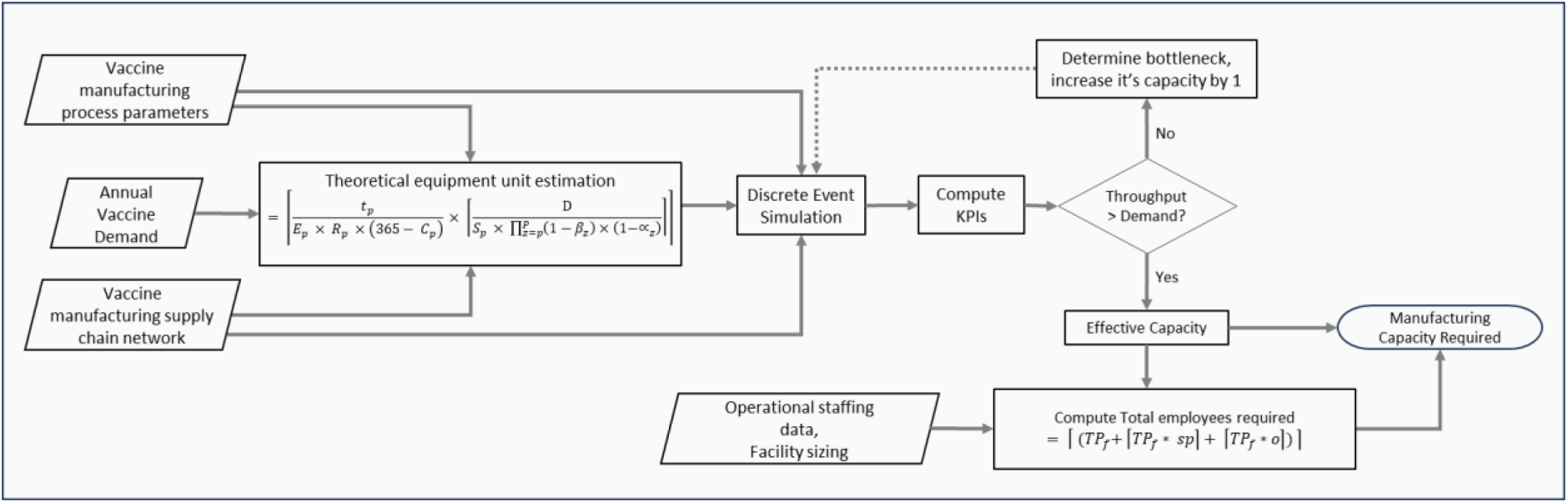
Modelling framework used to determine manufacturing capacity required to meet a target demand. Formulas are further explained in the subsection of calculating total equipment units and FTEs

**Figure 2:**
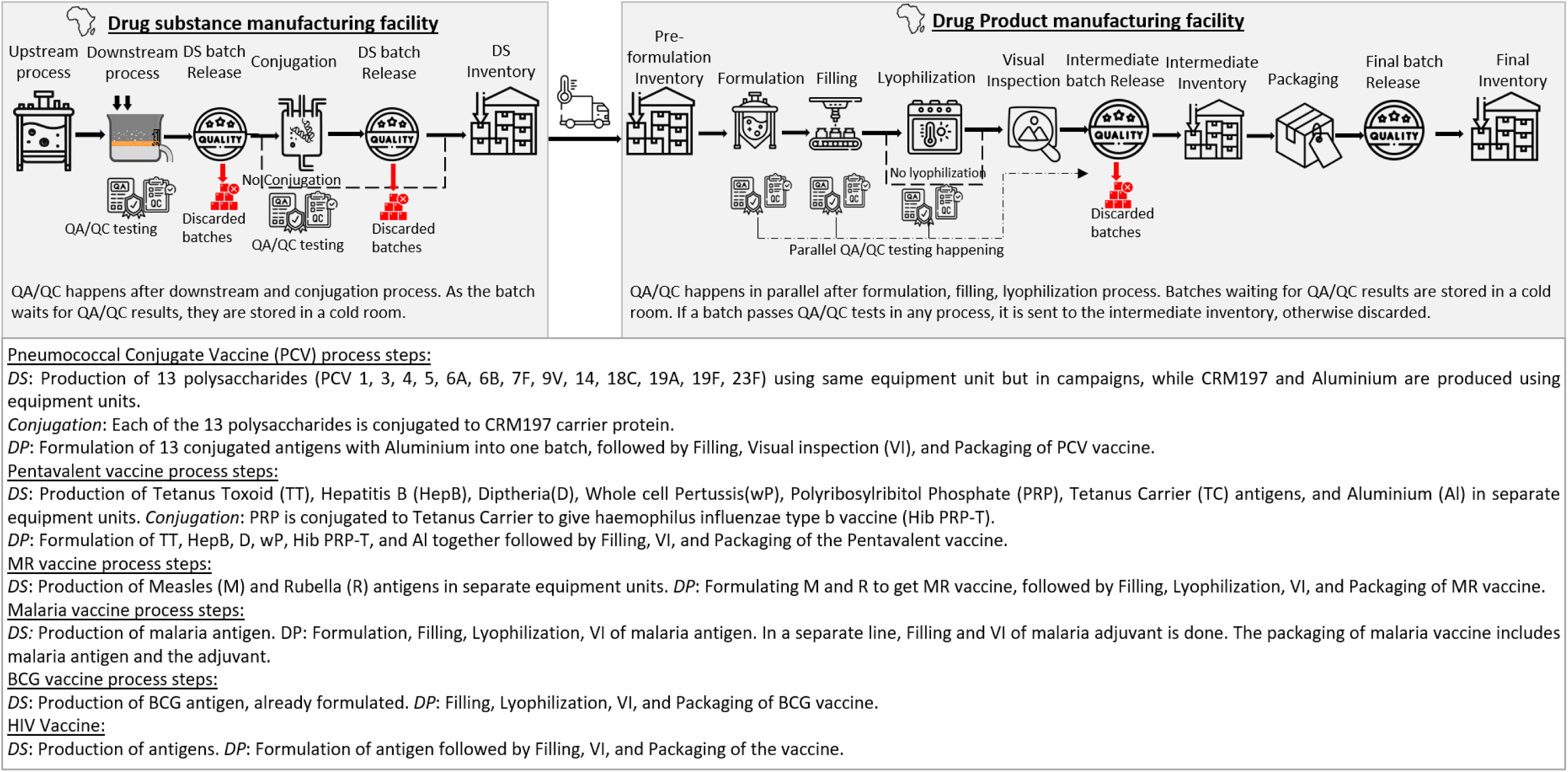
Visualisation of the generic structure of the VMSC network with additional description for PCV, Pentavalent, MR, Malaria, BCG, HIV, and COVID-19 vaccines. The VMSC network assumes that the drug substance is manufactured in a different facility from the drug product manufacturing facility. Both facilities are connected by a transportation module and if the transportation time is set to zero, it would resemble a VMSC with drug substance and drug product manufactured in the same facility. Icons are from https://www.flaticon.com/

### Step 1: Input data collection

#### Annual demand for selected vaccines

We used the annual vaccine demand target reported in Exhibit 15 and 16 of Africa’s Framework for Action ^1^ to estimate the DS and F&F manufacturing capacity required to fulfil the 2040 ambition. The required DS manufacturing capacity (in doses) for the prioritised vaccines are estimated at 139 million for Pentavalent, 90 million for MR, 88 million for Malaria, 82 million for HIV, 52 million for PCV, 52 million for BCG, 40 million for yellow fever, 266 million for COVID-19, and 12 million for Outbreak (Influenza, Chikungunya, Rift Valley fever, Lassa fever, and Disease X). For F&F production, the required capacities are 420 million for non-live virus vaccines, 260 million for live virus vaccines, and 130 million doses for viral vector vaccines. COVID-19 vaccine was excluded because it is not a priority, Yellow fever vaccine was excluded because of sufficient investment towards integrated manufacturing capacity at Institut Pasteur de Dakar ^26^, Outbreak vaccines were excluded because of their low aggregate demand, and no approved vaccines for routine use in Africa for Chikungunya, Rift Valley fever, Lassa fever, and Disease X.

#### Vaccine Manufacturing supply chain network

We consulted multiple subject matter experts (SME) with experience in vaccine manufacturing (see summary in Table 1) to design the most realistic VMSC network for each vaccine prioritised by the AfCDC. The VMSC network designed exclude research and development, raw materials, procurement, and distribution of vaccines from manufacturers to countries and vaccination points. The VMSC process steps that we modelled are shown in Figure 2 and provide an aggregate level representation of the VMSC. The process steps are broken down into DS production (upstream, downstream, conjugation - where applicable, QA/QC process) and F&F production (formulation, filling, lyophilization - where applicable, visual inspection (VI), packaging and QA/QC process), connected by a transportation module. The VMSC networks included in this study provide a wide range of complex VMSC networks. The generic networks include: F&F-only VMSC network (Live virus F&F, Non-live virus F&F, Viral vector F&F), single-antigen VMSC networks (HIV, BCG, and Malaria vaccine), multi-antigen VMSC networks with no sharing of equipment units among antigens (Pentavalent and MR vaccine), and multi-antigen VMSC networks with equipment units shared among antigens (PCV).

**Table 1:** Summary of phase 1 and phase 2 online meetings organized to collect vaccine manufacturing data from SMEs.

| Vaccine | Phase I |  |  | Phase 2 |  |
| --- | --- | --- | --- | --- | --- |
|  | # of meetings | # of Experts | # of organizations | # of meetings | # of Experts |
| Pentavalent: Tetanus toxoid, Tetanus Carrier, Diphtheria, wPertussis, Hepatitis B, PRP-T | 15 | 12 | 4 | 0 | 0 |
| MR | 2 | 4 | 2 | 2 | 3 |
| Covid-19 (data was used for HIV vaccine) | 3 | 7 | 2 | 2 | 1 |
| Pneumococcal | 2 | 6 | 2 | 2 | 3 |
| Tuberculosis | 2 | 5 | 2 | 2 | 1 |
| Malaria | 1 | 2 | 2 | 2 | 2 |
| Data validation during phase I | 5 | 5 | 2 | - | - |
| Total | 30 | 41 | 4 |  | 10 |

#### Manufacturing process parameters

We interviewed SMEs in two phases to collect data for the key vaccine manufacturing process parameters. During the first phase, we engaged 19 SMEs, affiliated to 4 different organisations, across 30 online meetings between June and July of 2022 (see Table 1). During these meetings, we collected VMSC assumptions and data on the manufacturing parameters. After all data were collected, five different validation meetings with a total of five experts were organised to validate the collected data for the different vaccine networks. In phase 2, we involved a different group of SMEs to validate the collected data in phase 1 using structured expert elicitation (SEE), following a modified IDEA protocol^27^. (see Appendix <u>A1.1</u>) This validation study was approved by the Social and Societal Ethics Committee (SMEC) of KU Leuven (G-2023-6398-R2). The validation sessions were carried out online with five DS and five F&F SMEs (see Table 1). For DS, some vaccines did not have any SME in the validation sessions, hence we used the DS manufacturing data collected during phase 1. Meanwhile, for F&F vaccines with no SMEs in validation study, DP manufacturing data of vaccines with similar manufacturing technologies collected in phase 2 were used. A limited number of SMEs participated in the validation because most of the vaccine specific SMEs had already contributed in phase 1 data collection and were ineligible for inclusion in this study. The manufacturing data collected and validated by SMEs is shown in Table 2 which includes batch size, processing times, and yield. QA/QC related data is shown in Table 3.

**Table 2:**
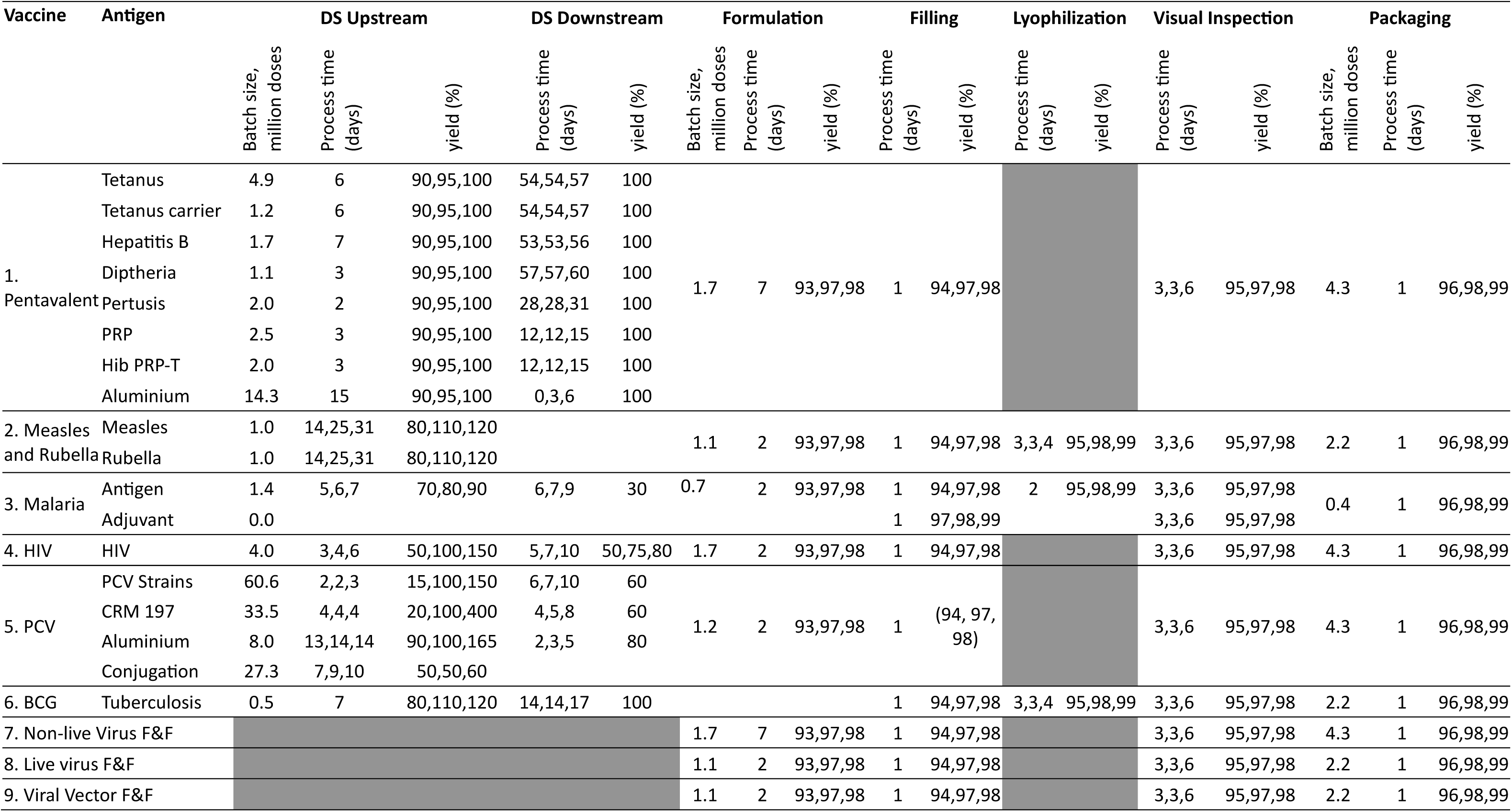
Key vaccine manufacturing process parameters used in the DES. Process times and yield are shown as minimum, most likely, maximum values a: For filling, we assume a filling speed of 250 vials/minute for Pentavalent, MR, PCV, BCG; 400 vials/minute for Malaria; 300 vials/minute for HIV. b: Lyophilization vial size (ISO) was assumed to be 10R for MR and BCG, 2R for Malaria. c: For Packaging, we assume 600 vials/minute for Pentavalent, HIV, and PCV; 300 vials/minute for MR, Malaria, and BCG.

**Table 3:**
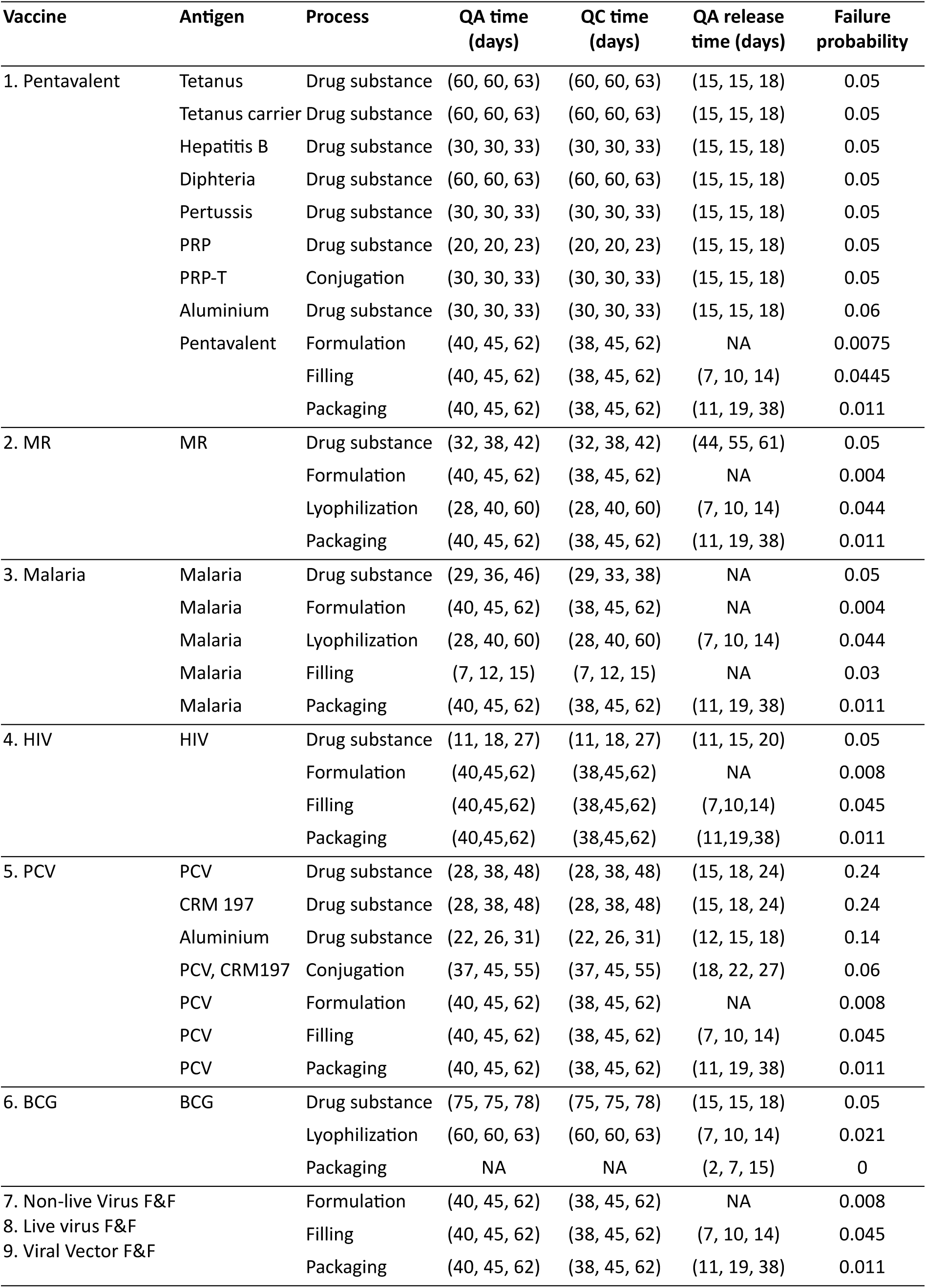
Input data for selected QA/QC parameters: testing time for Quality assurance (QA) and Quality control (QC), and total QA/QC failure probability for each vaccine and process.

### Step 2: Calculating total equipment units required for manufacturing using deterministic approach

We calculated the number of equipment units required at the *p^th^* process (*M_p_*) as the ratio of the total time (days) for the *p^th^* process to finish processing all batches within a year (*T_p_*) to the time each equipment unit in the *p^th^* process is available for batch processing annually (*A_p_*) ^22^. For any vaccine, antigen or adjuvant, *M_p_* can be formulated as:

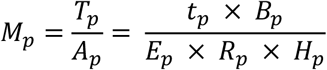

Where

- *p* = 1, 2, 3, 4, 5, 6, 7, 8, 9, corresponding to upstream, downstream, conjugation upstream, conjugation downstream, formulation, filling, lyophilization, visual inspection, and packaging process respectively
- *t_p_* is the most likely batch processing time (days) by an equipment unit in the *p^th^* process
- *E_p_* is the efficiency of the equipment units in the *p^th^* process (i.e. if the standard batch processing time is x days, will the equipment take exactly x days or longer)
- *H_p_* is the maximum total time available for an equipment unit in the *p^th^* process to process batches per year (days) i.e. 365 days – total days an equipment unit in the *p^th^* process is closed/unavailable for batch processing (*C_p_*).
- *R_p_* is the reliability (target utilisation) of the equipment units in the *p^th^* process, i.e. the proportion of maximum available time the equipment unit in the *p^th^* process will be busy processing batches in a year.
- *B_p_* is the total annual batches that must be processed by the equipment units in the *p^th^* process to meet the demand of the subsequent process (*p* + 1). The demand of the subsequent process (*p* + 1) adjusts the total annual demand *D*, for potential losses due to batch yield (*β*) and QA/QC failures (∝) in the *p^th^* process and all subsequent processes. This is done so that we start as many batches as possible to guarantee having enough batches to meet the target demand at the end of all processes ^25^. Let *S_p_* be the batch size, i.e. the total doses in a single batch in the *p^th^* process, 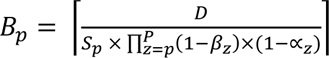 The total number of equipment units (*M_p_*) is therefore calculated as:

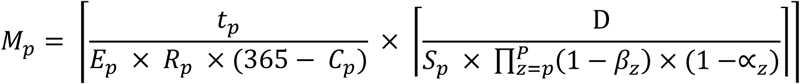

Based on stakeholder consultation, we assumed that *C_p_* is the sum of: 44 days allocated for weekly maintenance, 30 days for annual maintenance, 10 days for annual festivities/public holidays, and 14 days for media fill (filling process) or media form (formulation process). We further assumed that, as a new facility, the equipment units operate at 100% efficiency (*E_p_* = 1). For target utilisation, we first calculated total number of equipment units using utilisation target of 90% (i.e. *R_p_* = 0.9), followed by running the DES model using the calculated equipment units, and then evaluating the throughput. If the annual simulation throughput was less than demand, then a new calculation was done with an 80% utilisation target (i.e. *R_p_* = 0.8).

### Step 3: Simulating vaccine manufacturing process performance using DES

To simulate the vaccine manufacturing process shown in Figure 2, we applied a generic vaccine manufacturing DES model which has previously been used to simulate mRNA vaccine manufacturing^17^. This DES model was developed using Julia programming language ^28^ and provides a high degree of flexibility in rapidly testing a broad range of alternate VMSC networks and the impact of parameters such as process times, yields and QA/QC failure rates, on the VMSC network performance. During the simulation, the processing times, yields, and QA/QC test and retest durations for each batch are sampled from triangular distributions using the input data provided. The original DES model used a detailed structure for QA/QC testing, QA/QC personnel, and vaccine raw materials ^17^ but due to limited detailed level QA/QC data, we aggregated the detailed QA/QC structure in Kelchtermans et al ^17^ into three broad categories, namely QA testing, QC testing, and batch release testing (see Appendix Figure 6 for the structure). We also assumed unlimited availability of QA/QC personnel and raw materials.

For each VMSC network, we implemented a terminating DES, with the simulation starting on day zero and terminating after 5 years. We simulate the first five years of vaccine manufacturing to capture the production ramp-up period and evaluate the evolution of key performance indicators (KPIs) in a newly installed vaccine manufacturing facility. The simulation is initialized with no work-in-process or inventory and advances in daily time steps ^23^. Given the target demand and manufacturing process parameters elicited from SMEs, the model determines the number of batches required at the first manufacturing process by adjusting the target demand for expected yield losses and QA/QC failure probabilities across the production process ^25^. The batches are the main entity in the DES model entering the simulation at the DS upstream process and progressing through different manufacturing processes and tests as shown in Figure 2. Batches that fail any QA/QC retest are discarded and removed from the simulation. We assume sufficiently high demand at the start of the simulation such that the system remains capacity-constrained rather than demand-constrained, ensuring that bottleneck equipment is continuously supplied with batches for processing. Additionally, the model assumes a first-in/first-out batch queuing discipline and batch processing begins when an equipment unit becomes available and the remaining time before a planned equipment shutdown is greater than or equal to the maximum batch processing time.

#### Determining total replication for the DES model

We simulated multiple replications to account for uncertainty and variability inherent in the vaccine manufacturing processes. For each VMSC network, we determined the total number of replications (n) using a sequential procedure ^23^ in the following steps:

(i) We chose an initial total replications (*n* = *n*_0_ = 100) (> the recommended 10 replications^23^) and ran the DES model. ^14^
(ii) We calculated the average annual throughput in the fifth year, *X*(*n*), and the annual throughput precision, *δ*(*n*, *α*) of the simulation as

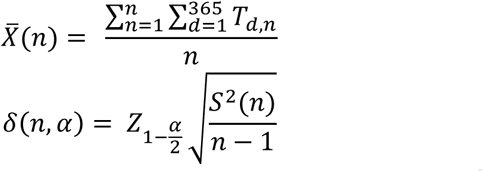 Where *n* is the number of replications, *T_d_*_,*n*_is the throughput for the *d^t^*^ℎ^ day and *n^t^*^ℎ^ replication, *α* is the significancy level (we assume 5%), 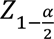 is the critical value, *S*^2^(*n*) is the variance of the throughput across the *n* replications.
(iii) We then calculated the ratio of the precision to the average annual throughput, i.e. 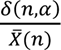

If the ratio was less than or equal to a prespecified relative error, we set the total replication required equal to the initial replication (*n*_0_), otherwise we increased the total replication and steps (i) to (iii) were repeated.

After running the DES model for each VMSC network 100 times, we observed that the relative error of the simulation average throughput in the fifth year ranged from 0.004 (BCG VMSC network) to 0.025 (PCV VMSC network) as shown in Table 4. These values are lower than the recommended threshold of 0.15 ^23^ hence total simulation replication is 100.

**Table 4:** Relative error of average annual throughput (5^th^ simulation year) of 100 replications of the base case DES model for different VMSC networks.

| VMSC Network | Total replications ($n$) | Average annual throughput ($\bar{X}(n)$) | Precision $\delta(n, \alpha)$ | $\frac{\delta(n, \alpha)}{\bar{X}(n)}$ |
| --- | --- | --- | --- | --- |
| Pentavalent | 100 | 151.0 | 0.852 | 0.006 |
| MR | 100 | 94.8 | 0.572 | 0.006 |
| Malaria | 100 | 91.6 | 0.365 | 0.004 |
| HIV | 100 | 118.5 | 0.968 | 0.008 |
| PCV | 100 | 56.7 | 1.390 | 0.025 |
| BCG | 100 | 51.9 | 0.216 | 0.004 |
| Non-live virus F&F | 100 | 530.7 | 1.560 | 0.003 |
| Live virus F&F | 100 | 356.0 | 1.120 | 0.003 |
| Viral vector F&F | 100 | 177.5 | 0.950 | 0.005 |

### Step 4: Calculating simulation KPIs

For each VMSC network simulated using the DES model, we calculated the average value and confidence intervals for two KPIs: annual throughput, and annual equipment utilisation. The computation of average and confidence interval of the KPIs are valid under the assumption that the replications are based on independent and identically distributed random variables ^23^. We applied the “fixed-sample-size procedure” described in ^23^ to calculate the confidence intervals of the KPIs. All analysis were done using R studio, version 2026.4.0.526 ^29^.

#### Annual throughput

Annual throughput was defined as the total number of vaccine doses released from the final inventory of a manufacturing facility in a particular year. We calculated the average total doses (*T̄_y_*), and the respective 95% confidence interval (*CIT_y_*) using the formula:

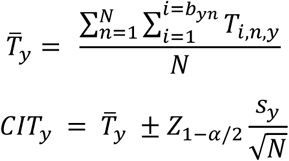

Where: *T̄_y_* is the average total throughput in the *y^t^*^ℎ^ year over all replications, *T_i_*_,*n*,*y*_is the total doses in the *i^t^*^ℎ^ batch released from the final inventory in the *y^t^*^ℎ^ year and *n^t^*^ℎ^ replication, *b_y_* is the number of batches released from the final inventory in the *y^t^*^ℎ^ year and *n^t^*^ℎ^ replication, *N* is the total number of replications, *CIT_y_* is the confidence interval of the average throughput of the *y^t^*^ℎ^ year, *s_y_* is standard deviation of the annual throughput in the *y^t^*^ℎ^ year, *α*=0.05.

#### Annual equipment utilisation

For each manufacturing process, we calculated the equipment utilisation as the ratio of the total time taken by all equipment units to process batches divided by the total time available for all equipment units to process batches within a particular year as shown below:

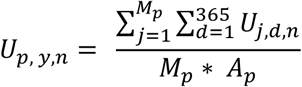

Where: *U_p_*_, *y*,*n*_ is the utilisation of all equipment units in the *p^th^* process, *y^t^*^ℎ^ year, and *n^t^*^ℎ^ replication; *U_j_*_,*d*,*n*_is the *j^t^*^ℎ^ equipment utilisation on the *d^t^*^ℎ^ day and *n^t^*^ℎ^ replication (note that *U_j_*_,*d*,*n*_= 1 if the equipment is utilized and zero otherwise); *M_p_* is the number of equipment units in the *p^th^* process ; *A_p_* is the time each equipment unit in the *p^th^* process is available for batch processing annually.

Given *U_p_*_, *y*,*n*_, we calculated the average yearly utilisation (*Ū_p,y_*) and confidence interval of the annual utilisation for the *p^th^* process and *y^t^*^ℎ^ year (*CIU_p_*_,*y*_) as:

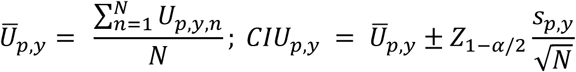

Where: *Z*_1−*α*/2_ is the z value for the 95% confidence level, *s_p_*_,*y*_ is the standard deviation of average equipment units utilisation for the *p^th^* process and *y^t^*^ℎ^ year over all replications.

### Step 5: Determining available manufacturing capacity to fulfil the target demand

We compared the DES model’s average annual throughput to the annual demand to ascertain if demand was fulfilled. If the average annual throughput was greater than or equal to target demand for that given year, we concluded that the capacity in the VMSC is sufficient. However, if the average annual throughput was less than annual demand, we determined the bottleneck process by checking the equipment units with the highest utilisation and increase its total by one ^25^. We reran the DES model with the updated equipment units, and the average annual throughput was evaluated again. This was repeated until we found the total equipment units that led to an average annual throughput greater than or equal to the target demand. This total number of equipment units that fulfils demand was then called the effective equipment units.

### Step 6: Total number of facilities and fulltime employees required

Under facility sizing, we first determined which vaccines can be manufactured in the same facilities. We opted for a mix of DS-only, integrated DS and F&F, and F&F-only manufacturing facilities. In addition, we also accounted for vaccine technology, as well as vaccine type, contamination risks, and vaccine presentations, when combining multiple antigens or vaccines in the same facility. For each manufacturing facility, we calculated the total FTEss required as the sum of technical employees, technical support employees, and administrative employees adjusted for absenteeism. Total technical employees in facility *f* (*TP_f_*) was calculated as the sum of the product of total parallel equipment units, number of technical employees per shift, total number of shifts per working day, and absenteeism adjustment over all manufacturing processes. Meanwhile, we calculated the total personnel providing technical and administrative support as a proportion of the total technical personnel as shown below:

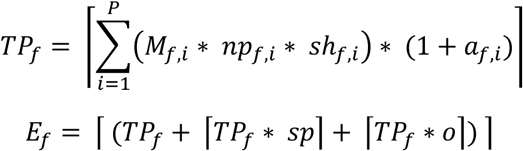

Where: the total number of fulltime employees required in facility *f* is *E_f_*, the number of equipment units for facility *f* required at process *i* is *M_f_*_,*i*_, total number of personnels required per shift in facility *f* and process *i* is *np_f_*_,*i*_, total number of shifts per day in facility *f* and process *i* is *s*ℎ*_f_*_,*i*_, percentage adjustment for absenteeism for facility *f* and process *i* is *a_f_*_,*i*_, percentage adjustment for technical support personnel is *sp*, and percentage adjustment for administrative employees is *o*. The operational staffing data, including shifts per day, absenteeism per work shift, additional support personnel, and administrative personnel were collected from SMEs (see Appendix Table 10 for data).

### Scenario testing

#### Base case

The base case scenario simulates the VMSC network using the calculated total equipment units and parameter values collected from the SMEs. We further assume that the DS and DP are in the same manufacturing locations and hence the transportation time from the DS to F&F facility is zero.

#### Simulation Scenarios

In addition to the base case, we analysed ten different scenarios shown Table 5. In each scenario, we varied only one parameter and kept the VMSC network and other parameters the same as in base case. The first two scenarios explored the impact of reduced batch yield at DS upstream and DP formulation process on manufacturing capacity. These were motivated by AstraZeneca’s manufacturing partners experiencing varying batch yields, with some sites producing three times more vaccine doses from a single batch than low productive sites ^30^The third scenario explored how a 50% reduction in batch size for all manufacturing processes impacts manufacturing capacity. Batch size can reduce because research and development might show that a higher antigen concentration per dose may be preferred if it provides higher immune responses and vaccine efficacy. For example, a study evaluated different dose levels of hemagglutinin in influenza vaccines (i.e. 15µg, 45µg, and 135µg) and found that the higher dose levels elicited higher antibody levels after vaccination^31^. Hence with the same manufacturing equipment size, less doses can be realised.

**Table 5:** Overview of the simulation scenarios used to evaluate DS and DP manufacturing capacity.

| Scenario | Scenario name | Explanation |
| --- | --- | --- |
| 1 | DS Half Yield | The base case yield values (minimum, most likely, and maximum) for the DS upstream manufacturing process are reduced by 50%. |
| 2 | DP Half Yield | The base case yield values (minimum, most likely, and maximum) for the formulation process are reduced by 50%. |
| 3 | Half Batch Size | The base case batch size for all the manufacturing process is reduced by 50% due to increase in antigen concentration per dose (dose size). |
| 4 | DS Time +1 day | The processing time for DS upstream manufacturing process is increased by 1 day. |
| 5 | DP Time +1 day | The processing time for each of the DP processes (Formulation, Filling, Lyophilization, VI, Packaging) is increased by 1 day. |
| 6 | DS Unavailable +30 days | The total number of days that equipment units in the DS upstream process are unavailable for batch processing is increased by 30 days. |
| 7 | DP Unavailable +30 days | The total number of days that equipment units in the DP Formulation process are unavailable for batch processing is increased by 30 days. |
| 8 | Doubling DS QA/QC deviation probability | The batch QA/QC deviation probability for DS is doubled. |
| 9 | Doubling DP QA/QC deviation probability | The batch QA/QC deviation probability for DP processes are doubled. |
| 10 | Transport 1 Week | The transportation time of batches from DS site to F&F site ranges from 5 to 7 days. |

The fourth and fifth scenario explored the impact of a 1-day increase in batch processing time for both DS and DP processes, due to additional cleaning requirements, batch changeover activities, or slower cleaning performance due to limited staffing on capacity. We also explored scenarios on the impact of increasing the time of DS and DP equipment unavailability by 30 days in the sixth and seventh scenario, representing unplanned manufacturing equipment failure not accounted for in the base case DES model. The impact of doubling the QA/QC deviation probability at both DS and DP facilities, which may arise from personnel errors and differences in staff experience levels ^32^, was also evaluated in the eighth and ninth scenario. Lastly, we evaluated the impact of manufacturing DS and DP in separate facilities, with a transportation time of 5–7 days from the DS to the DP site since base-case assumes DS and DP are manufactured within the same facility with zero transportation time.

We applied the paired-t confidence interval approach described in ^23^ to test for the difference in the scenarios average annual throughput and the base case average annual throughput. To account for multiple comparisons across the ten scenarios evaluated, the significance level was adjusted using Bonferroni correction (i.e. α = 0.05/10). If the confidence interval (CI) of the difference included zero, the scenario was considered not significantly different from base case.

## Results

We used the deterministic method to estimate the total number of equipment units required to meet target demand for each vaccine using VMSC data collected from SMEs. Initially, we assumed a target utilisation of 90% and all vaccines, except for MR and Pentavalent vaccines, had higher annual throughput compared to demand (Table 6). For MR and Pentavalent vaccines, annual throughput did not meet demand, and the target utilisation was reduced to 80% for selected antigens (MR, Tetanus toxoid, Diptheria, and PRP), which resulted in a unit increase in the total equipment units. Overall, the estimated equipment units required were 66 for DS upstream, 16 for formulation, 12 for filling, 10 for lyophilization, and 9 for packaging process. The theoretical total number of equipment units were used as input to the DES model to simulate the base case for each of the VMSC network. The resulting 95% confidence interval (CI) of the annual throughput in the fifth simulation year shows that there is enough capacity to meet the annual vaccine demand for all the vaccines since the CI includes the target annual demand (see Table 6).

**Table 6:**
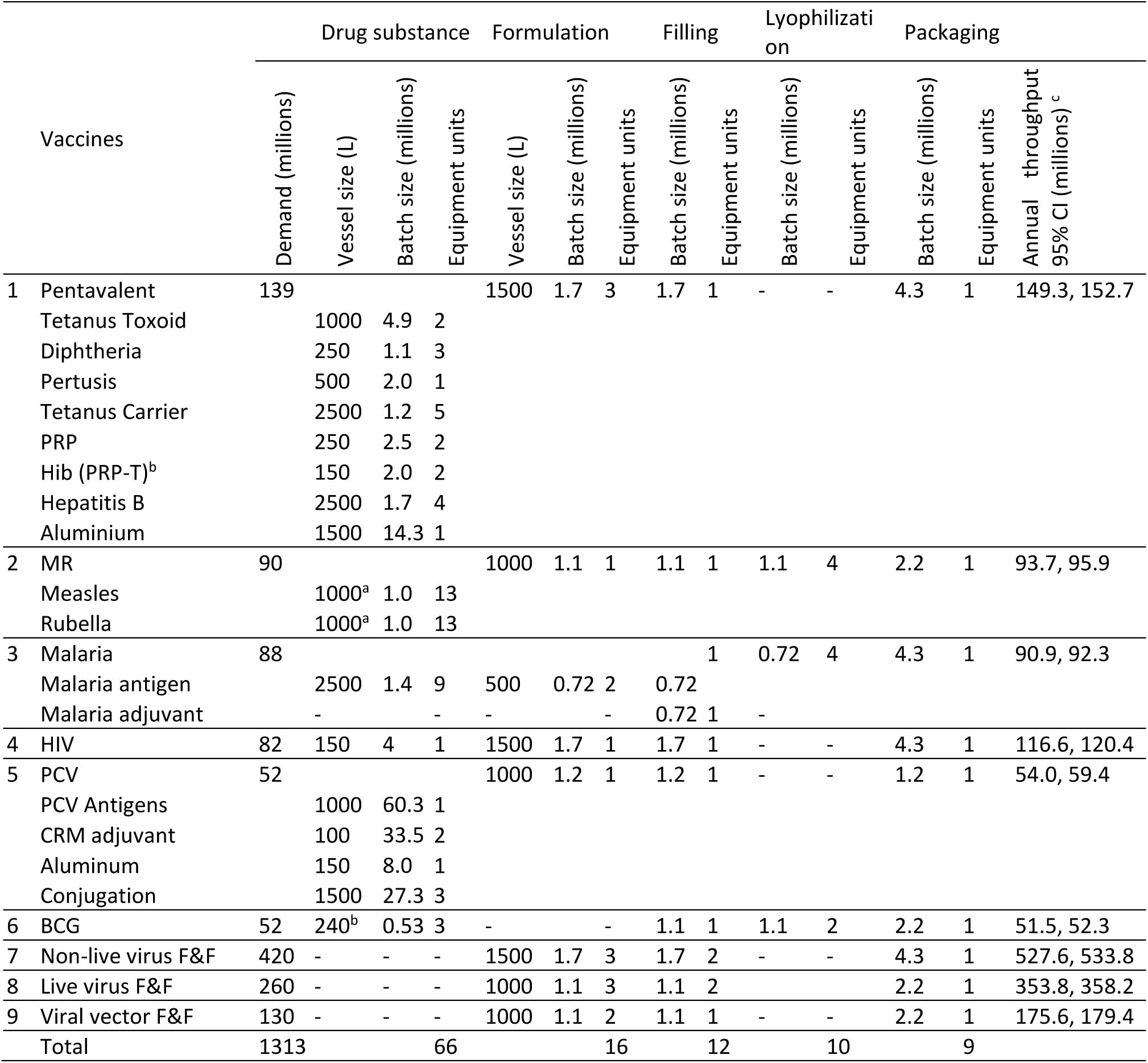
Number of equipment units required in different manufacturing processes to meet target demand for specific vaccines. ^a^ We assume 100 roller bottles each of size 100 ml to make 1000L. ^b^ We assume 8 parallel 30L vessels to make 240L for BCG. ^c^ CI is confidence interval (lower confidence interval, upper confidence interval) of the fifth year of simulation.

Using the number of equipment units shown in Table 6 as input in the DES model, Figure 3 shows the time the first batch is seized and released from different processes under base case conditions assuming production begins on 1^st^ of January. The first packaged vaccine doses are released from the final inventory in the seventh month for Malaria, BCG and HIV vaccine, end of the eighth month for MR vaccine, and towards the end of 12^th^ month for Pentavalent vaccines (Figure 3). For all vaccines, QA/QC testing takes the most considerable amount of time where vaccines must wait for tests results before proceeding to the next step. For multivalent vaccines like Pentavalent, formulation only begins after all the required antigens have enough released DS batches. For example, the first batch of PRP antigen finishes the DS release test around the 50^th^ day but must wait for the Tetanus Carrier antigen which is released on day 138 before it can start the conjugation process, followed by QA/QC testing and release ending on day 200. This means that the aluminium adjuvant and whole cell Pertussis antigens, which had DS batch release on day 63 and day 76, respectively, have to wait until day 208 for formulation of all the antigens to begin (see Figure 3).

**Figure 3:**
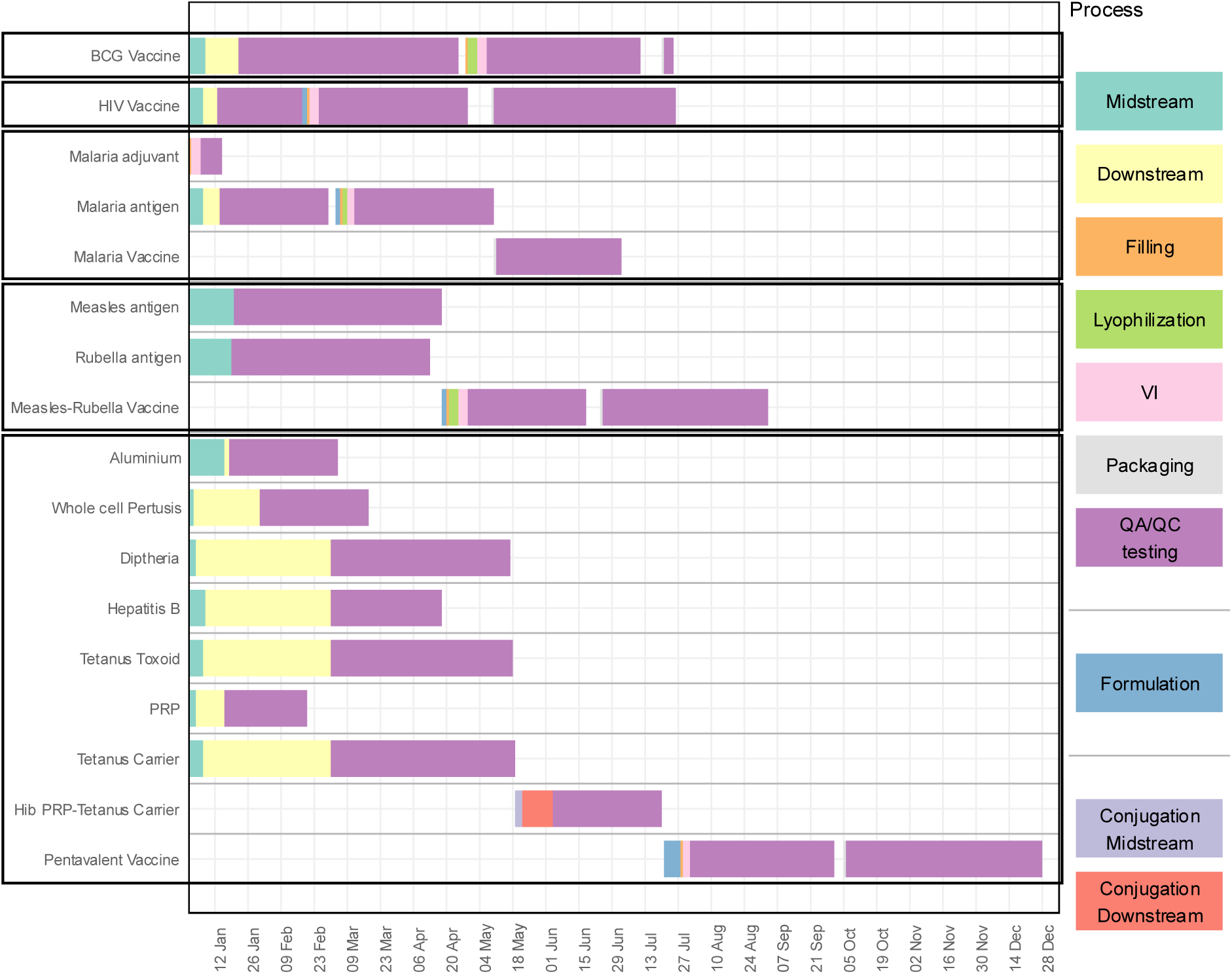
Visualization of how the time the first batch enters and leaves the different manufacturing processes and QA/QC testing for different vaccines. VI means Visual Inspection. And QA/QC testing includes time spend for QA, QC, and batch release.

The cumulative monthly throughput for the different vaccines in each simulation year is shown in Figure 4. The proportion of target annual demand fulfilled in the first year for each vaccine was estimated at: 50.4% for BCG, 62.6% for HIV, 50.2% for Malaria, 21.9% for MR, 0% for PCV, 4.5% for Pentavalent vaccine, 79.9% for Live virus F&F, 74.5% for Non-Live virus F&F, and 79.0% for Viral vector F&F. The cumulative annual throughput for BCG, HIV, Malaria, and Pentavalent vaccines matched demand in the second year of manufacturing compared to MR and PCV vaccines whose demands were matched in the third year of manufacturing (Figure 4). The intersection of the base case cumulative annual plot with the demand (Figure 4) indicates that, for HIV vaccine, demand is already met by the 8^th^ month of the second year, whereas for all other vaccines, the demand is only fulfilled by the end of the simulation year. A comparison of the CI of the cumulative annual throughput further shows that the PCV vaccine has a wider CI compared to other vaccines (Figure 4 e). This is because PCV has a highly variable DS batch yield compared to other vaccines.

**Figure 4:**
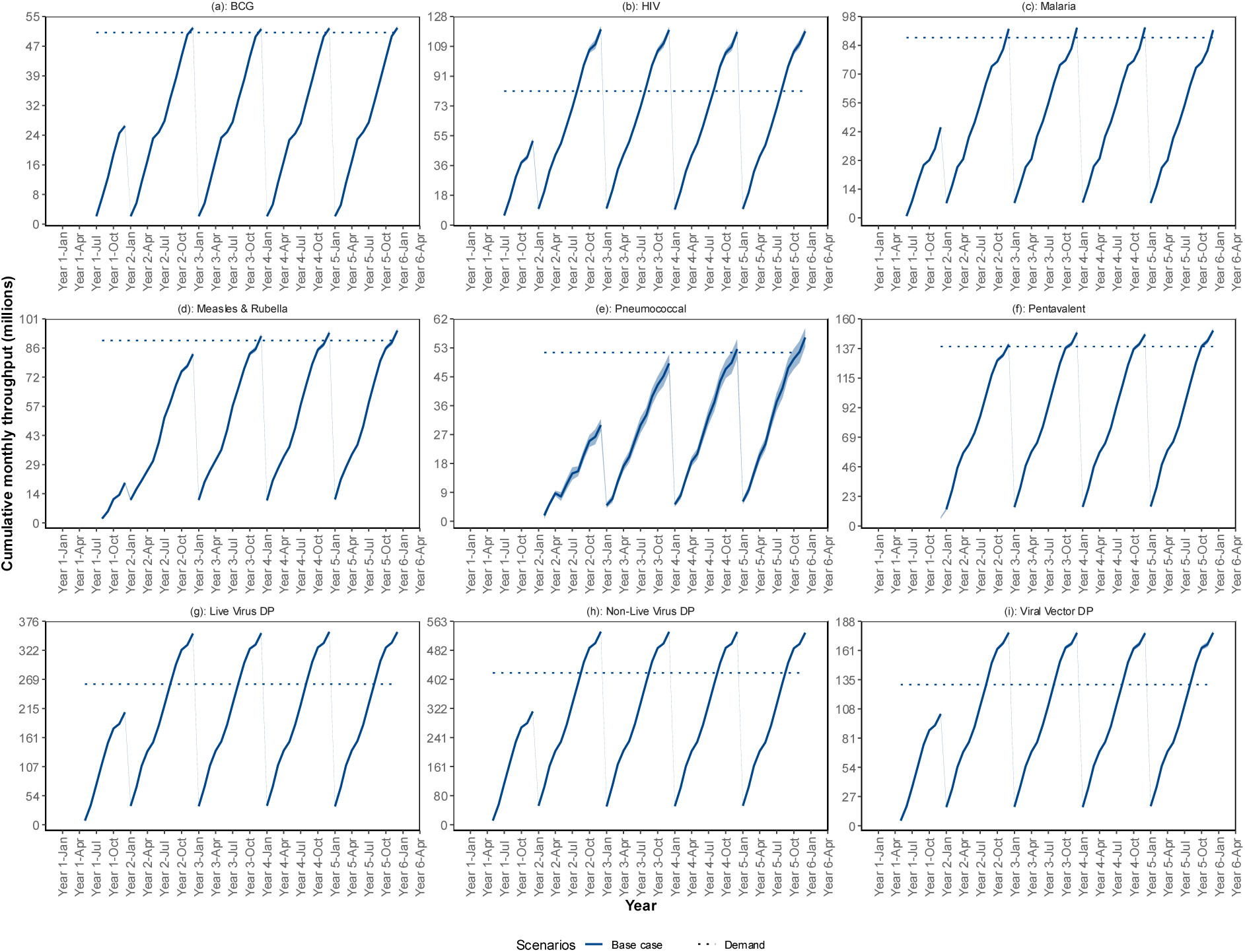
Visualization of the base case cumulative monthly throughput for different vaccines (showed per year) relative to their respective annual demand, visualized by the dotted line. The curves represent the average across 100 simulation replications for each month, while the light blue shaded region represents the 95% confidence interval around the average cumulative throughput for each year. Each vaccine uses a different y- axis scale to improve visibility since the annual demand varies across vaccines

We constructed a paired-t confidence interval to assess whether there were significant differences between the alternative scenarios and base case (see Table 7). The results show that for all vaccines there was no significant difference between base case and the scenario of having one week transportation time from DS to DP. There was no significant difference in throughput when increasing the DP facility unavailability by 30 days compared to the base case for HIV, Malaria, PCV, and MR vaccines. Meanwhile increasing the DS facility unavailability by 30 days leads to a substantial reduction in the throughput compared to base case for all vaccines (see Table 7).

**Table 7:** Confidence intervals (CIs) for the difference in average annual throughput between scenarios and the base case for vaccines with integrated DS and F&F VMSC network. Values in brackets denote the lower and upper CI bounds (million doses). Bonferroni adjustment was applied for multiple comparisons (α=0.05/10). Confidence intervals including zero shows that there is no significant difference and they are coloured red. ^a^ for BCG, since there is no formulation process, scenario 7 was not explored.

| Scenario | Scenarios name | BCG | HIV | Malaria | Measles & Rubella | Pneumococcal | Pentavalent |
| --- | --- | --- | --- | --- | --- | --- | --- |
| 1 | DS Half Yield | [-26.5, -25.3] | [-61.8, -55.8] | [-46.4, -44.2] | [-49.9, -46.3] | [-32.2, -24.2] | [-75.4, -69.8] |
| 2 | DP Half Yield | [-26.5, -25.3] | [-62.2, -56.4] | [-46.6, -44.4] | [-49.8, -46.4] | [-31.1, -23.1] | [-79.1, -73.9] |
| 3 | Half Batch Size | [-26.7, -25.3] | [-62.7, -57.1] | [-46.5, -44.5] | [-49.8, -46.6] | [-31.6, -24] | [-74.2, -69] |
| 4 | DS Time +1day | [-8.1, -6.5] | [-25.9, -19.1] | [-14.3, -11.7] | [-7.4, -3.4] | [-24.8, -14.8] | [-36.9, -31.3] |
| 5 | DP Time +1day | [-0.9, 0.5] | [-5.6, 1.2] | [-16.5, -14.5] | [-23.6, -20] | [-5.6, 5] | [-24.4, -18] |
| 6 | DS Unavailable +30days | [-7.5, -6.1] | [-15.9, -9.5] | [-10.2, -7.8] | [-15.3, -11.1] | [-10.2, -0.2] | [-14.1, -8.1] |
| 7 | DP Unavailable +30days | NA<sup>a</sup> | [-2.8, 4] | [-0.6, 1.8] | [-2.6, 1.2] | [-5, 4.6] | [-23.7, -17.7] |
| 8 | Doubling DS QA/QC deviation probability | [-4.5, -2.9] | [-10.5, -3.9] | [-6.1, -3.5] | [-9.7, -5.9] | [-36.3, -26.7] | [-7.3, -1.1] |
| 9 | Doubling DP QA/QC deviation probability | [-2.7, -1.3] | [-20.8, -14.4] | [-14.7, -11.7] | [-17.6, -13.4] | [-16.7, -5.7] | [-28.9, -22.5] |
| 10 | Transport 1 Week | [-1.1, 0.3] | [-3.5, 3.3] | [-0.5, 1.9] | [-2.6, 1.6] | [-8.2, 2.2] | [-4.7, 0.7] |

Increasing DS downstream process time by one day led to a significant reduction in throughput compared to base case for all vaccines, with the CI of reduction in millions of doses being highest for Pentavalent vaccines [-36.9, -31.3] and lowest for Measles & Rubella vaccines [-7.4, -3.4]. Increasing the batch processing times in DP processes by one day had no significant effect on throughput for BCG, HIV, and PCV vaccines (Table 7). This is because the base case utilisation levels of the DP equipment units were relatively low, such that the increase in processing times did not cause a bottleneck and, therefore, had an insignificant effect on throughput (Figure 5). In contrast, a significant reduction in throughput was observed for Malaria (-16.5, -14.5), MR (-23.6, -20), and Pentavalent (-24.4, -18) million doses (Table 7). For Malaria and MR vaccines, this was driven by an increase in the average formulation equipment utilisation, from 63% to 95%, and from 85% to 98%, respectively (Figure 5). For Pentavalent vaccine, the formulation process was already a bottleneck in the base case, with an average capacity utilisation of 97%. Hence increasing DP processing time by one day further shifted the manufacturing pressure by increasing average filling equipment utilisation from 33% to 56%, and in packaging process from 10% to 18% (Figure 5). For vaccines such as HIV, BCG, and PCV, although equipment utilisation increased in the extended DP processing time scenario, the effect was not enough to cause significant change in the annual throughput compared to base case (see Figure 5 and Table 7).

**Figure 5:**
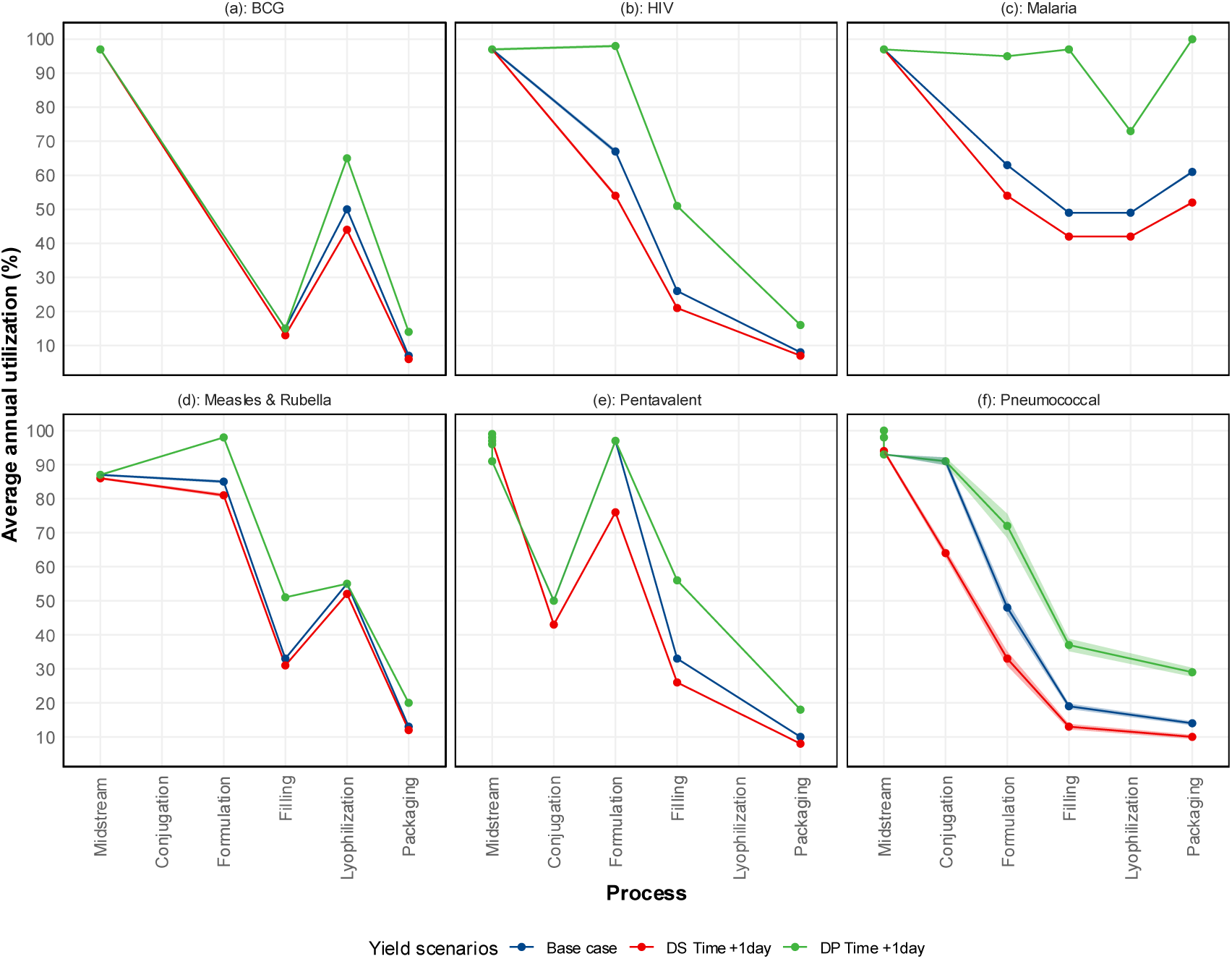
Average annual utilisation of equipment units in the fifth simulation year under different vaccines

For all vaccines, halving the batch size, and reducing DS yield and DP yield by 50%, significantly reduced the annual throughput compared to the base case (see Table 7). For these three scenarios, comparing the ratio of each scenario’s lower CI of the difference (Table 7) and annual vaccine demand (Table 6) shows that HIV vaccine had the highest reduction with annual throughput approximately 75% below annual demand, followed by the PCV vaccine at about 60% below demand, while the BCG vaccine shows the smallest reduction at approximately 51% below annual demand (see Table 7). Similarly, doubling the QA/QC deviation probability in both DS and DP processes significantly reduced throughput across all vaccines relative to the base case. For HIV, Malaria, Measles & Rubella, and Pentavalent vaccines, doubling the DP QA/QC deviation probability has a greater impact on reducing average annual throughput; at least twice the reduction observed when the deviation probability is doubled in DS processes only. In contrast, for PCV and BCG vaccines, doubling the DS QA/QC deviation probability results in a larger reduction in average annual throughput than doubling the deviation probability in DP processes (Table 7). These scenarios reveal the impact of process variability on production throughput, allowing for iterative estimation of manufacturing capacity requirements under different real-world conditions.

Eleven manufacturing facilities would be required in Africa to meet demand. Five of these would be integrated DS and F&F facilities, two would be DS-only facilities, and four would be F&F-only facilities (see Table 8). Three out of the eleven manufacturing facilities would be dedicated for Pentavalent vaccines. One DS-only facility includes a spore-forming suite for Tetanus toxoid antigen, and a yeast cell-based culture suite for producing Hepatitis B, Diphtheria, and Pertussis antigens. Another DS-only facility produces Tetanus carrier in a separate suite from Polyribosylribitol Phosphate (PRP). The pentavalent F&F-only facility is where PRP and tetanus carrier conjugation and fill-finish of the vaccine occurs. The five integrated DS and F&F facilities would be for PCV (a conjugated non-live virus vaccines), BCG (live virus, mycobacterium and shake flasks used in DS), HIV (mRNA vaccine, DS uses IVT reactor), MR (live attenuated vaccine, DS uses roller bottle technology), Malaria (Protein based technology, DS uses Bioreactor).

**Table 8:** Total FTEs required per vaccine manufacturing site. ^a^ the DS and DP total technical staff adjusts the personnel required to operate the DS lines in all shifts for absenteeism; ^b^ Total support staff is assumed to be 100% of total technical staff; ^c^ Total administrative staff is calculated as 15% of the sum of total technical and support staff; ^d^ Total FTEs is the sum of total technical, support, and administrative staff. To avoid having total staffs in decimal numbers after multiplication, we take the ceiling of the respective values for e.g. 217.2 becomes 218.

| Site # | Antigens Manufactured | DS Technical staffs <sup>a</sup> | DP Technical staffs <sup>a</sup> | Total technical staffs | Total Support staffs <sup>b</sup> | Total administrative staffs <sup>c</sup> | Total FTEs <sup>d</sup> |
| --- | --- | --- | --- | --- | --- | --- | --- |
| 1 | DS-only: Tetanus, Diphtheria, Pertussis, and Hepatitis B | 600 | 0 | 600 | 600 | 180 | 1380 |
| 2 | DS-only: TT Carrier, PRP | 513 | 0 | 513 | 513 | 154 | 1180 |
| 3 | F&F-only: Pentavalent PRP-T Conjugation | 44 | 211 | 255 | 255 | 77 | 587 |
| 4 | Integrated DS-DP: MR | 312 | 197 | 509 | 509 | 153 | 1171 |
| 5 | Integrated DS-DP: Malaria | 540 | 184 | 724 | 724 | 218 | 1666 |
| 6 | Integrated DS-DP: HIV | 29 | 103 | 132 | 132 | 40 | 304 |
| 7 | Integrated DS-DP: PCV | 262 | 129 | 391 | 391 | 118 | 900 |
| 8 | Integrated DS-DP: BCG | 36 | 114 | 150 | 150 | 45 | 345 |
| 9 | F&F-only: Viral Vector F&F | 0 | 117 | 117 | 117 | 36 | 270 |
| 10 | F&F-only: Live virus F&F | 0 | 173 | 173 | 173 | 52 | 398 |
| 11 | F&F-only: Non-Live Virus F&F | 0 | 173 | 173 | 173 | 52 | 398 |
|  | Total | 2336 | 1401 | 3737 | 3737 | 1125 | 8599 |
|  | % total FTEs | 27% | 16% | 43% | 43% | 13% |  |

A total of 8,599 FTEs would be required of which about 43% (3,737) would be DS and DP technical staff, 43% (3,737) would be support staff (including warehouse, resource planning, QC/QA and engineering staff), and 13% (1,125) would be administrative staff (see Table 8). The integrated facility for Malaria vaccine would be the largest with an estimated 1,666 FTEs while the viral vector F&F-only facility would be the smallest with 270 FTEs.

## Discussions and Conclusion

This study presented a systematic approach to estimating vaccine manufacturing capacity requirements by integrating deterministic capacity estimation methods with discrete event simulation to account for vaccine manufacturing complexities, uncertainties and variabilities. The deterministic approach used demand, batch size, batch processing times, batch yield, QA/QC deviation probabilities, annual equipment availability, machine efficiency, and target utilisation as input parameters. As such, it provides a simplified estimation tool for decision makers and manufacturing facility planners, particularly when simulation expertise, computational resources, or detailed manufacturing data are limited.

However, our results show that this approach can lead to optimistic capacity estimates because variability and uncertainty in key process parameters are not explicitly considered. For example, the deterministic method suggests that demand can be met with fewer equipment units under a 90% target utilisation level, whereas the simulation results indicate that for vaccines with smaller batch sizes and higher variability in processing times and yields, yearly demand is not consistently reached in all the simulation years. In contrast, a target utilisation of 80% provides additional buffer capacity through slightly higher equipment units, enabling all vaccine networks to meet demand even when there are high variability and delays in upstream manufacturing processes ^11^. In addition, having excess capacity provides greater flexibility to manufacture other vaccines, and provide a buffer in case of disruptions, as well as unanticipated spikes in demand observed during outbreaks ^33^. However, expanding capacity by increasing total equipment units instead of alternatives like reducing the unavailability of equipment units, improving manufacturing process quality, or reducing equipment setup and cleaning time ^12^, and investing in process intensification ^34^, requires substantial investments. If capacity is not fully utilised, it could be difficult for manufacturers to attain competitive vaccine prices and returns on investment critical for long-term financial sustainability because of relatively high manufacturing costs ^35^.

Our simulated scenarios show that a reduction in the DS and DP process yield significantly decreases throughput for all the vaccine networks, with no substantial difference in magnitude when the yield reduction occurs in DS or DP process. This indicates that yield losses at any stage will limit final output because they directly determine how many doses can proceed to the next manufacturing step. This finding aligns with factory physics principles ^25^, and biopharmaceutical studies showing that lower-than-expected titres can substantially increase the required manufacturing capacity, leading to external contract manufacturing or the construction of new manufacturing facilities to maintain supply ^36^. Yield reductions also have important economic implications, as the same raw materials, staff, fixed infrastructure and operating resources are used to produce fewer doses ^37^ Overall, these findings demonstrate that maintaining stable and predictable process yields is critical for a sustainable manufacturing capacity that meets required vaccine demand in Africa.

Our study further demonstrates that the effect of an increase in batch quality deviations on manufacturing capacity is substantial, and depends on the vaccine and the supply chain stage. Quality deviations increases lead time and variability because of the extra time spent investigating whether the deviation is substantial enough for the batch to be rejected, hence the reduction in the expected annual throughput ^25^. For instance, our simulation showed that doubling of DS-related quality deviations led to a greater reduction in annual throughput compared to doubling of DP-related quality deviations for PCV and BCG vaccines. This is because the extra quality checks and retests of batches further delays the start of DP processes ^10^, more especially for PCV vaccine with 13 antigens produced in campaigns using one equipment. Other vaccines like HIV, Malaria, MR, and Pentavalent vaccines had enough DS batches in their inventory which made the impact of deviation in the DS-related process to be considerably lower compared to DP-related deviations. This points to the importance of having buffer capacity and released DS vaccine batches in inventory, what factory physics calls finished good inventory ^25^. Furthermore, quality does not impact only throughput, but also facility uptime. A simulation study showed that in integrated upstream and downstream production trains, an increase in the number of downstream processes and failure rates leads to a reduction in the facility uptime ^38^. Hence, our findings reinforce the importance, when evaluating vaccine manufacturing capacity requirements, of considering not only the frequency of quality deviations, but also the manufacturing process in which they occur, buffer capacity and inventory.

Vaccine manufacturing capacity estimation becomes even more complex for multivalent vaccines as it depends not only on the individual manufacturing process capacity but also on several interacting system level factors. This system level capacity is influenced by several factors. Firstly, the individual antigen manufacturing process variability is amplified when multiple antigens are combined into one vaccine ^4^. For example, one study showed that the theoretical probability of performing a potency retest increases from 5% to about 40% in the case when 10 antigens are combined into one vaccine. Therefore, the possibility of having a delay in formulating the multiple antigens increases since it is more likely for at least one antigen to fail a batch release testing ^39^. Secondly, capacity is not only determined by the number of equipment units required for each antigen, but also by whether the timing and quantity of released DS batches for all antigens are well synchronised. For Pentavalent vaccine for example, Whole Cell Pertussis antigen batches must wait for over 4 months for the slowest antigen (Hib-PRPT) before formulation can start. Similar behaviour has been observed in assembly systems, where throughput is constrained by the last-arriving component in converging flows, creating waiting times that reduce overall system output ^40^. Thirdly, it introduces the need for not only additional storage requirements but also validating the storage duration within which the DS or intermediate products would still be stable for further conjugation or formulation of all antigens ^41^. Studies have shown that when components in converging flows do not arrive simultaneously, waiting times increase, which directly translates into higher intermediate stock, since components must be stored under conditions until all required inputs are available for assembly ^25,40^. Hence when no action is taken to reduce manufacturing processes variability and improve batch scheduling, non-synchronization arises which increases batch waiting time and buildup of inventory in the facility, and further limiting manufacturing capacity ^25^. In this context, DES is helpful because it explicitly captures and shows how delays and variability in one antigen affect the timing of final multivalent vaccine throughput, hence informing proper intermediate storage capacity needs.

Vaccine manufacturers ought to ensure predictable and reliable process yields, and manufacturing quality which is more likely if skilled and experienced staff are available ^4^. We estimated that about 3,737 DS and DP technical staff are required in Africa, yet only about 1,600 vaccine manufacturing staff are available in Africa as of 2022 according to AfCDC ^1^. Hence there is need for considerable investments towards training, recruiting and retaining future vaccine manufacturing experts in the continent ^20,42^. For existing technical and non-technical staffs, their skills and experience should be improved through continuous reskilling, practical courses and apprenticeships ^43,44^, to ensure availability of highly skilled and experienced staff capable of not only producing consistently high quality vaccines, but maintaining less variable batch yield and processing time, low and less frequent equipment breakdown, and better production planning.

This study, being a DES, is limited by the assumptions made and the quality of the input data. Our approach to data collection and validation with stakeholders provide simulation input data with some degree of quality. Secondly, our determination of total facilities and FTEs required did not take costs into account, which greatly determines how big or small a facility should be. In addition, our current method does not account for factors that can influence vaccine demand and supply such as: product characteristics ^45^, vaccine costs, reimbursement levels and quality ^46^, general public’s attitudes, behaviour and acceptability of vaccines ^47^, trust in locally made vaccines ^48^, uncertainty in vaccine price and demand in the presence of competition from other vaccine manufacturers ^4,49^. These factors are all interdependent and can generate feedback loops that can complicate long term capacity planning and investments. A systems perspective using stakeholder-led systems dynamics modelling ^50,51^ offers a way to integrate vaccine demand and supply dynamics ^52^ and could be extended to support long-term vaccine manufacturing capacity planning. Therefore, in subsequent studies, simulation models could embed the VMSC in a broader vaccine value chain or vaccine manufacturing eco-system, that integrates all these factors influencing vaccine demand and supply.

In conclusion, this study demonstrated the need for substantial investments in vaccine manufacturing in Africa in order to realise the AfCDC 2040 ambitions for a local and sustainable vaccine supply. Those investments must be coordinated and extended across different vaccines, vaccine manufacturing platforms and technologies in the continent to avoid overcapacity in a few vaccine technologies and platform as witnessed with the massive investments in mRNA technology post-COVID-19 and undercapacity in others ^5,21^. Furthermore, strengthening R&D capabilities and investments, as well as technology transfer efforts, is key to promote the long-term manufacturing of vaccines that aligns with regional priorities ^53,54^. Our capacity estimation approach can provide valuable evidence to support the coordination of manufacturing infrastructure establishment and expansion, workforce development strategies, assess the feasibility of future manufacturing capacity before major investments are made, and align manufacturing capacity with market demand, which are key working streams within the AfCDC framework for action ^1^.

## Data availability statement

All relevant vaccine manufacturing process data are provided in the manuscript. Because of confidentiality, detailed QA/QC data can only be made available from the authors upon approval from both KU Leuven and PATH.

## Acknowledgements

This work was funded by PATH, via a grant from the Gates Foundation GAT.586257_01720152-CRT. The conclusions and opinions expressed in this work are those of the author(s) alone and shall not be attributed to the Foundation. Under the grant conditions of the Foundation, a Creative Commons Attribution 4.0 License has already been assigned to the Author Accepted Manuscript version that might arise from this submission. Please note works submitted as a preprint have not undergone a peer review process

We would like to thank all the subject matter experts from PATH, CEPI, AfCDC, and other confidential organisations for their invaluable contributions through the provision of data, experiences, and constructive feedback throughout the project. We are also grateful to Raafat Fahim, Vishal Mukund Sonje, Valentijn Stienen and Leonor Guariguata for their valuable comments and suggestions. Finally, we also acknowledge Marika Engel whose reflections on vaccine manufacturing modelling while at Access-To-Medicines provided us with rich modelling lessons.

## Author information

*Access-To-Medicines Research Centre, Faculty of Economics and Business, KU Leuven:* Junior Ocira, Donovan Guttieres, Robin Kelchtermans, Carla Van Riet, Lise Boey, Nico Vandaele

*Operations Management Research Group, KU Leuven:* Nico Vandaele

*International Dispensary Association:* Eki Ramnaps Eghosasere

*NRL Enterprise Solutions:* Kristopher Howard

*PATH:* Mauro Bernuzzi, Steven D. Dong, Jens Demand, Simone Blayer

*Università Cattolica del Sacro Cuore. Faculty of Economics. Milano (Italy):* Mauro Bernuzzi

## Contributions

**Funding acquisition:** N.V, S.B, J.D. **Project administration:** S.D.D, N.V, S.B, L.B. **Conceptualisation:** J.O, N.V, S.B, J.D, M.B. **Data Collection and Validation:** J.O, D.G, E.R, C.V.R, L.B, K.H, M.B, S.D.D, J.D, S.B, N.V. **Methodology:** J.O, D.G, R.K, E.R, C.V.R, L.B, K.H, M.B, J.D, S.B, N.V., **Software:** R.K, **Data analysis:** J.O, N.V., **Supervision:** N.V, S.B, J.D, S.D.D, M.B, L.B, C.V.R. **Writing original draft:** J.O, N.V, S.B, M.B. **Writing review & editing:** J.O, D.G, R.K, E.R, C.V.R, L.B, K.H, M.B, S.D.D, J.D, S.B, N.V;

## Ethics declarations

The authors declare no competing interests

## Use of Large Language Models (LLMs)

The authors declare that ChatGPT (OpenAI) was used for copy editing of the manuscript texts and debugging R code during the analysis of simulation output. All decisions regarding background, methodology, analysis, interpretation of results, discussion and conclusions were made by the Authors.

## Appendix

### A1.1 Phase 2 vaccine manufacturing data collection using structured expert elicitation IDEA protocol

To enhance the scientific validity of the data provided by subject matter experts, we adopted a structured expert elicitation method to validate the manufacturing data collected in phase 1 of the study. We applied a slight modification of the IDEA protocol ^27^, which is a structured expert elicitation method for collecting experts estimates. The method derives its name from its four steps: Investigate, Discuss, Estimate and Aggregate. The choice of the IDEA protocol was informed by its ease of use, relatable questions format, and suitability in accommodating time and resource constraints while still achieving the project’s aim. We collect the minimum, maximum and most likely estimates for the variables of interest from multiple experts using the four-questions format, and aggregates these estimates to obtain the final estimates. The four questions format additionally asks experts to provide the degree of belief for the estimates provided that reflects experts level of confidence that the real value of the variable falls within the minimum and maximum. The method has been shown to reduce overconfidence in interval judgments and help experts construct quantitative estimates ^27^. As a result of participants time and availability constraints, participants requested to discuss proposed aggregated estimates rather than to send in independent round 2 estimates, hence the modification to the IDEA protocol. We show an example of the data collection and analysis template in Table *9*.

**Table 9:** Structured expert elicitation data extraction and analysis template.

|  | Round 1 estimates |  |  |  |  |  |  |  | Round 2 estimates |  |  |  |  |  |  |
| --- | --- | --- | --- | --- | --- | --- | --- | --- | --- | --- | --- | --- | --- | --- | --- |
|  | Experts estimates |  |  |  | Standardized estimates |  |  |  | Experts estimates |  |  |  | Standardized estimates |  |  |
|  | Min | Max | Best | Confidence | Min | Max | Best |  | Min | Max | Best | Confidence | Min | Max | Best |
| Expert 1 |  |  |  |  |  |  |  |  |  |  |  |  |  |  |  |
| Expert 2 |  |  |  |  |  |  |  |  |  |  |  |  |  |  |  |
| Min |  |  |  |  |  |  |  |  |  |  |  |  |  |  |  |
| Max |  |  |  |  |  |  |  |  |  |  |  |  |  |  |  |
| Mean |  |  |  |  |  |  |  |  |  |  |  |  |  |  |  |

**Table 10:** Parameter values used to calculate total fulltime employees (FTEs) per facility.

| # | Facility | DS |  | Conjugation |  | Formulation |  | Filling (& Lyophilization) |  | Filling diluent |  | Visual Inspection |  | Packaging |  |
| --- | --- | --- | --- | --- | --- | --- | --- | --- | --- | --- | --- | --- | --- | --- | --- |
|  |  | #Personel/<br>equipment | #Shifts/day | #Personel/<br>equipment | #Shifts/day | #Personel/<br>equipment | #Shifts/day | #Personel/<br>equipment | #Shifts/day | #Personel/<br>equipment | #Shifts/day | #Personel/<br>equipment | #Shifts/day | #Personel/<br>equipment | #Shifts/day |
| 1 | DS-only: Tetanus, Diphtheria, Pertussis, and Hepatitis B | 22 | 2.25 |  |  |  |  |  |  |  |  |  |  |  |  |
| 2 | DS-only: TT Carrier, PRP | 27 | 2.25 |  |  |  |  |  |  |  |  |  |  |  |  |
| 3 | F&F-only: Pentavalent PRP-T Conjugation |  |  | 9 | 2 | 15 | 2.25 | 15 | 2.25 |  |  | 13 | 2 | 6 | 2 |
| 4 | Integrated DS-DP: MR | 4 | 2.50 |  |  | 5 | 2.5 | 20 | 2.25 | 15 | 2.25 | 7 | 2 | 6 | 2 |
| 5 | Integrated DS-DP: Malaria | 22 | 2.25 |  |  | 5 | 2.25 | 15 | 2.25 | 15 | 2.25 | 7 | 2 | 6 | 2 |
| 6 | Integrated DS-DP: HIV | 12 | 2.00 |  |  | 5 | 2.25 | 15 | 2.25 |  |  | 13 | 2 | 6 | 2 |
| 7 | Integrated DS-DP: PCV | 27 | 1.50 | 9 | 2 | 15 | 2.25 | 15 | 2.25 |  |  | 13 | 2 | 6 | 2 |
| 8 | Integrated DS-DP: BCG | 10 | 1.00 |  |  | 5 | 2.25 | 15 | 2.25 | 15 | 2.25 | 7 | 2 | 6 | 2 |
| 9 | F&F-only: Viral Vector F&F |  |  |  |  | 5 | 2.25 | 15 | 2.25 |  |  | 13 | 2 | 6 | 2 |
| 10 | F&F-only: Live virus F&F |  |  |  |  | 5 | 2.25 | 15 | 2.25 |  |  | 13 | 2 | 6 | 2 |
| 11 | F&F-only: Non-Live Virus F&F |  |  |  |  | 5 | 2.25 | 15 | 2.25 |  |  | 13 | 2 | 6 | 2 |
|  | Adjustment for absenteeism | 20% |  |  |  |  |  |  |  |  |  |  |  |  |  |
|  | Adjustment for support personnel | 100% |  |  |  |  |  |  |  |  |  |  |  |  |  |
|  | Adjust for administrative personnel | 15% |  |  |  |  |  |  |  |  |  |  |  |  |  |

Since our simulation assumes parameters follows triangular distribution, we asked experts to provide minimum, most likely and maximum values. These inputs were then standardized using the IDEA protocol’s formula that standardizes the experts inputs to a prespecified credible interval (we assume 90% credible intervals) to obtain the uncertainties of all experts across questions on a consistent scale^27^. The formula used is shown below.

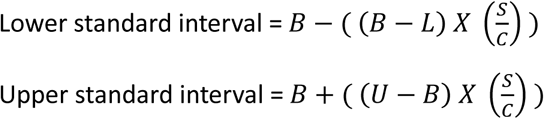

Where: *B* is the best guess, *L* is the minimum estimate, *U* is the maximum estimate, *S* is the target credible interval level to standardized estimates to, and *C* is the level of confidence given by participant. Below we show the total number of experts who were interviewed per vaccine.

**Figure 6.**
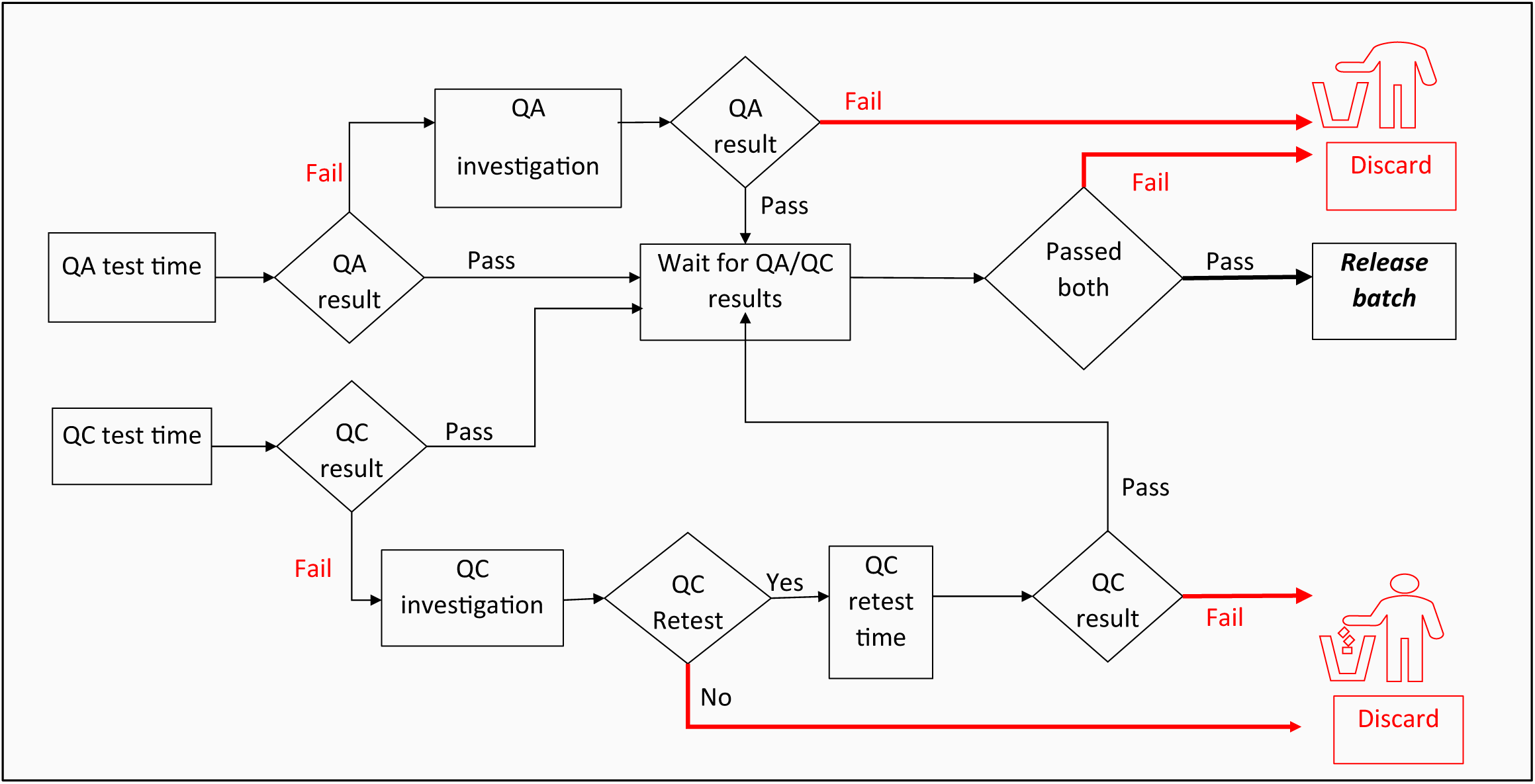
Batch: Quality assurance (QA) and Quality Control (QC) testing process implemented in the simulation model. Each batch is tested and only batches that pass both QA and QC are released, while those that fail either QA or QC are discarded

## Notes

### Competing Interest Statement

The authors have declared no competing interest.

### Author Declarations

This validation study was approved by the Social and Societal Ethics Committee (SMEC) of KU Leuven (G-2023-6398-R2).

